# Cross-sectional associations between lifecourse dementia risk factors, plasma neurodegenerative biomarkers, and cognition in older adults in India

**DOI:** 10.1101/2025.09.19.25336183

**Authors:** Sarah Gao, Alden L. Gross, Leon M. Aksman, Masroor Anwar, Eileen M. Crimmins, Sharmistha Dey, Abhishek Gupta, Bharat Thyagarajan, Jinkook Lee, Emma Nichols

## Abstract

**BACKGROUND:** Plasma neurodegenerative biomarkers are a potential low-cost tool for studying Alzheimer’s disease and dementia in population-based research, especially in low- and middle-income countries (LMIC). However, their associations with modifiable risk factors and utility as an outcome in epidemiologic studies remain unclear.

**OBJECTIVE:** Our objective was to estimate the cross-sectional association between modifiable lifecourse risk factors for dementia and plasma-based neurodegenerative biomarkers, and to compare those with the associations between lifecourse risk factors and cognition in a population-representative Indian sample.

**METHODS:** Using nationally representative data from the Longitudinal Aging Study in India-Diagnostic Assessment of Dementia (N=1625, average age 68.2 years), we estimated linear regressions to compare cross-sectional associations between lifecourse risk factors and both neurodegenerative biomarkers (β-amyloid 42/40, total-tau, phosphorylated Tau181, GFAP, NfL) and cognitive outcomes (general cognition, memory).

**RESULTS:** Despite significant associations between seven of thirteen risk factors and cognitive outcomes, associations between risk factors and neurodegenerative biomarkers were largely null with some exceptions; for example, hypertension (β=0.17SD; 95% CI:0.08,0.26) and diabetes (β=0.21SD; 95% CI:0.09, 0.32) were associated with higher NfL.

**CONCLUSIONS:** While we found expected associations between lifecourse risk factors for dementia and cognition, there was not strong evidence of cross-sectional associations between risk factors for dementia and plasma-based biomarkers.

## Background

As public health and economic conditions have improved globally, low- and middle-income countries (LMIC) are experiencing a rapid demographic shift with a greater proportion of the population living to old age^1,2^ and increasing burden from Alzheimer’s disease and related dementias (AD/ADRD) (hereafter referred to as dementia).^3,4^ It is especially pertinent to study dementia in India, as India is currently the most populous country in the world^5^ and its population is exposed to risk factors unique from high-income countries, where most dementia research is concentrated.^6,7^ Despite its salience, studying dementia is challenging in India. In dementia epidemiology more generally, it is often cost-prohibitive to ascertain clinical dementia status, leading many studies to use continuous measures of cognition as alternative outcomes given that impairment in cognition is one of the two key pieces of a dementia diagnosis.^8^ However, in India and other contexts with high levels of illiteracy and innumeracy, many cognitive tests are difficult to administer.^9,10^ Differences in language and culture pose challenges in adapting cognitive tests.^10,11^ In contrast, research using biomarkers to study factors associated with the biological underpinnings of dementia may be less susceptible to challenges due to the adaptation and administration of cognitive tests. However, a lack of infrastructure and resources has complicated efforts to measure biological mechanisms or specific age-related processes through conventional methods such PET and MRI.^6^

New breakthroughs in measuring plasma neurodegenerative biomarkers may address these issues. Blood-based neurodegenerative biomarkers are easier and less expensive to collect on a large scale than other biomarker modalities, such as PET imaging and cerebrospinal fluid collection, and they have been shown to distinguish stages of AD and detect AD pathology.^12–15^

However, research on these biomarkers has primarily been conducted in clinical settings^13–15^ or population-based studies in high-income countries,^15,16^ and more work is needed to characterize blood-based neurodegenerative biomarkers in LMICs. Prior analyses have shown that biomarkers are associated with cognition across a range of settings, including India.^17–20^ However, observed effect sizes have been small and relatively few studies in LMIC settings have used these biomarkers in further investigation. Thus, better understanding of the associations between modifiable risk factors and plasma neurodegenerative biomarkers would provide important insights into the utility of plasma neurodegenerative markers as outcomes in epidemiologic research on dementia, particularly in LMICs.

In this study, we used nationally representative data from the Longitudinal Study in India – Diagnostic Assessment of Dementia (LASI-DAD) to compare associations of established dementia risk factors and cognitive outcomes versus associations between the same risk factors and blood-based neurodegenerative biomarkers. Given the evidence of small effect sizes linking biomarkers and cognitive outcomes, we hypothesize that there will be differences in observed associations between risk factors and the two sets of outcomes.

## Methods

### Sample

We used data from Wave 1 (2017-2019) of the LASI-DAD study, an ongoing study of dementia in older adults aged 60+ in India.^21^ LASI-DAD is the only nationally representative study of cognition and aging in India, drawing data from 14 states across the country.^22,23^ LASI-DAD is a sub-study of the broader nationally representative Longitudinal Aging Study in India (LASI), which uses a multistage stratified area probability cluster sampling design to ensure representativeness at both the national level and for each state and union territory.^22^ Of the 4,096 respondents from Wave 1, we restricted our analytic sample to participants who had complete data on neurodegenerative biomarkers and cognitive testing (N=1,625). The distribution of demographic and socio-economic characteristics is similar between included and excluded respondents (Appendix Table 1). The LASI-DAD study was approved by the University of Southern California IRB (approval no. UP-15-00684) on September 28, 2016.

### Blood-based neurodegenerative biomarkers

Fasting venous blood samples (17 mL) were collected from participants by trained phlebotomists. The samples were transported to a local lab for initial processing; if the lab was more than two hours away from the study site, an onsite automatic centrifuge machine was available to process blood samples. Following initial processing, separated plasma samples for neurodegenerative biomarker assays were shipped via cold chain at -20°C to the Metropolis Healthcare Ltd laboratory, New Delhi, and stored at -80°C until further analysis. Assays for β-amyloid 42 (Aβ42), β-amyloid 40 (Aβ40), total tau, phosphorylated tau 181 (pTau-181), glial fibrillary acidic protein (GFAP), and neurofilament light (NfL) were conducted at the Department of Biophysics, All India Institute of Medical Sciences (AIIMS), New Delhi, using an ultrasensitive and automated Single Molecule Array (Simoa) analyzer (Quanterix HD-X). We used the ratio of β-amyloid 42 (Aβ_42_) and β-amyloid 40 (Aβ_40_) to quantify amyloid pathology.

All biomarker variables were log-transformed and z-standardized to a mean of 0 and a standard deviation of 1.

### Cognitive test measures

Participants completed a comprehensive battery of cognitive tests adapted from the Harmonized Cognitive Assessment Protocol^22,23^ to accommodate differences in language, literacy, and culture.^19^ The battery included tests covering diverse cognitive domains, including orientation, memory, language/fluency, executive functioning, and visuospatial functioning (full list in the Appendix).” Confirmatory factor analysis was used to develop estimates of latent cognitive functioning, as well as domain-specific functioning.^24^ These factor scores were scaled to a mean of 0 and a standard deviation of 1 within the LASI-DAD Wave 1 sample. We used estimates of general cognitive functioning and memory in this analysis, as memory is typically most closely related to AD pathology.^25,26^

### Lifecourse dementia risk factors

We used all lifecourse dementia risk factors established by the 2024 *Lancet* Commission on dementia except traumatic brain injury, which was not captured in the LASI-DAD Wave 1 survey instrument.^27,28^ These risk factors include: educational attainment, hypertension, diabetes, high cholesterol, obesity, vision impairment, hearing impairment, depression, social isolation, physical inactivity, excessive alcohol consumption, smoking, and air pollution (details in Appendix). The social isolation index measure was z-standardized to increase comparability and interpretability.

### Other variables

Age, sex/gender, educational attainment, marital status, and caste were self-reported. Socioeconomic status was determined by per-capita household consumption (quintiles).

Body-mass index (BMI) was calculated based on measured height and weight. Estimated Glomerular Filtration Rate (eGFR) was calculated using the CKD-EPI creatinine-cystatin C equation^29^ to measure chronic kidney disease.

### Statistical analysis

We used descriptive statistics to characterize the analytic sample. We used linear regression to estimate cross-sectional associations between dementia risk factors and neurodegenerative biomarkers, and between dementia risk factors and cognitive function.

All models were adjusted for age and sex/gender with additional adjustments to control for potential confounding depending on the outcome. Cognition models were additionally adjusted for education, household consumption, marital status, and caste. Biomarker models were additionally adjusted for BMI and eGFR given prior evidence suggesting that BMI and kidney function may impact the measurement of blood biomarkers.^30^ Cognition models where education was the exposure dropped education from the adjustment set and biomarker models where BMI the exposure dropped BMI from the adjustment set. To assess whether additional adjustment for socio-economic status indicators in cognition models impacted comparisons, we conducted a sensitivity analysis with additional adjustment for education, household consumption, marital status, and caste in the biomarker models. Additionally, we performed sensitivity analysis of all models stratified by Clinical Dementia Rating (CDR) to analyze heterogeneity by dementia disease stage. Online consensus diagnosis was used in the LASI-DAD sample to generate the following diagnoses: cognitively normal, mild cognitive impairment or questionable dementia (referred to as MCI), and dementia.^31^

## Results

### Cohort characteristics and descriptive results

Descriptive statistics of the analytic sample (n=1,625) are shown in Table 1. Mean age was 68.7 years (SD=7.2), and approximately 47% of the sample was women. Nearly half of respondents (49%) reported no formal schooling. The most common caste classification was other backwards class (41.8%), followed by no caste or other caste (35.2%), scheduled class (19.1%), and scheduled tribe (3.9%).

**Table 1.** Characteristics of the LASI-DAD W1 sample with complete data on dementia risk factors, cognitive test measures, and blood-based neurodegenerative biomarkers (n=1625)

| Variable | Mean (SD) or % (N) |
| --- | --- |
| Age (years) | 68.7 (7.2) |
| Female | 47.1 (765) |
| Education |  |
| No schooling | 48.6 (790) |
| Less than secondary | 27.3 (444) |
| Secondary or higher | 24.1 (391) |
| Per capita consumption quintile |  |
| 1 (lowest) | 18.6 (302) |
| 2 | 21.8 (354) |
| 3 | 20.8 (338) |
| 4 | 19.9 (323) |
| 5 (highest) | 19.0 (308) |
| Caste |  |
| Scheduled caste | 19.1 (311) |
| Scheduled tribe | 3.8 (62) |
| Other backward class | 41.8 (680) |
| No caste or other caste | 35.2 (572) |
| Married | 67.4 (1096) |
| eGFR (mL/min) | 71.6 (19.9) |
| Hypertension | 61.9 (1006) |
| Diabetes | 18.2 (295) |
| High cholesterol | 26.8 (436) |
| BMI |  |
| Underweight (<18) | 17.7 (287) |
| Normal (18.0-22.9) | 41.9 (681) |
| Overweight (23.0-24.9) | 14.0 (227) |
| Obese (≥25) | 26.5 (430) |
| Visual impairment |  |
| No visual impairment | 47.8 (777) |
| Mild visual impairment | 16.8 (273) |
| Moderate visual impairment | 32.3 (525) |
| Severe visual impairment/blind | 3.1 (50) |
| Hearing problem | 10.8 (176) |
| CIDI depression score (0-7) | 0.5 (1.6) |
| Social isolation score (0-12) | 6.4 (1.9) |
| Physical inactivity | 33.5 (545) |
| Excessive alcohol consumption | 1.8 (30) |
| Currently smoking | 23.0 (374) |
| Unclean cooking fuel | 43.2 (702) |
| GFAP | 152.7 (103.4) |
| NfL | 45.0 (93.7) |
| ptau181 | 42.2 (25.9) |
| Total tau | 2.6 (3.8) |
| Aβ ratio | 0.1 (0.1) |
| General cognition | 0.0 (0.9) |
| Memory | 0.0 (0.9) |

### Associations between risk factors and biomarkers

We did not observe a clear pattern of strong or statistically significant associations between risk factors and neurodegenerative biomarkers (Figure 1). However, we identified a few associations of note. We observed small but significant positive associations between multiple risk factors for cardiovascular disease and elevated NfL. Specifically, hypertension (β=0.17SD, p <0.001) and diabetes (β=0.21SD, p <0.001) were associated with increased NfL; observed effects were equivalent to approximately 5 (blood pressure) or 6 (HbA1c) years of age. NfL was also marginally associated with several other dementia risk factors, including underweight BMI and moderate vision impairment. Vision impairment was associated with lower Aβ_42/40_ ratio, indicative of higher amyloid pathology in the brain. Associations were statistically significant for the moderate vision impairment category and were marginally significant in the mild category.

**Figure 1.**
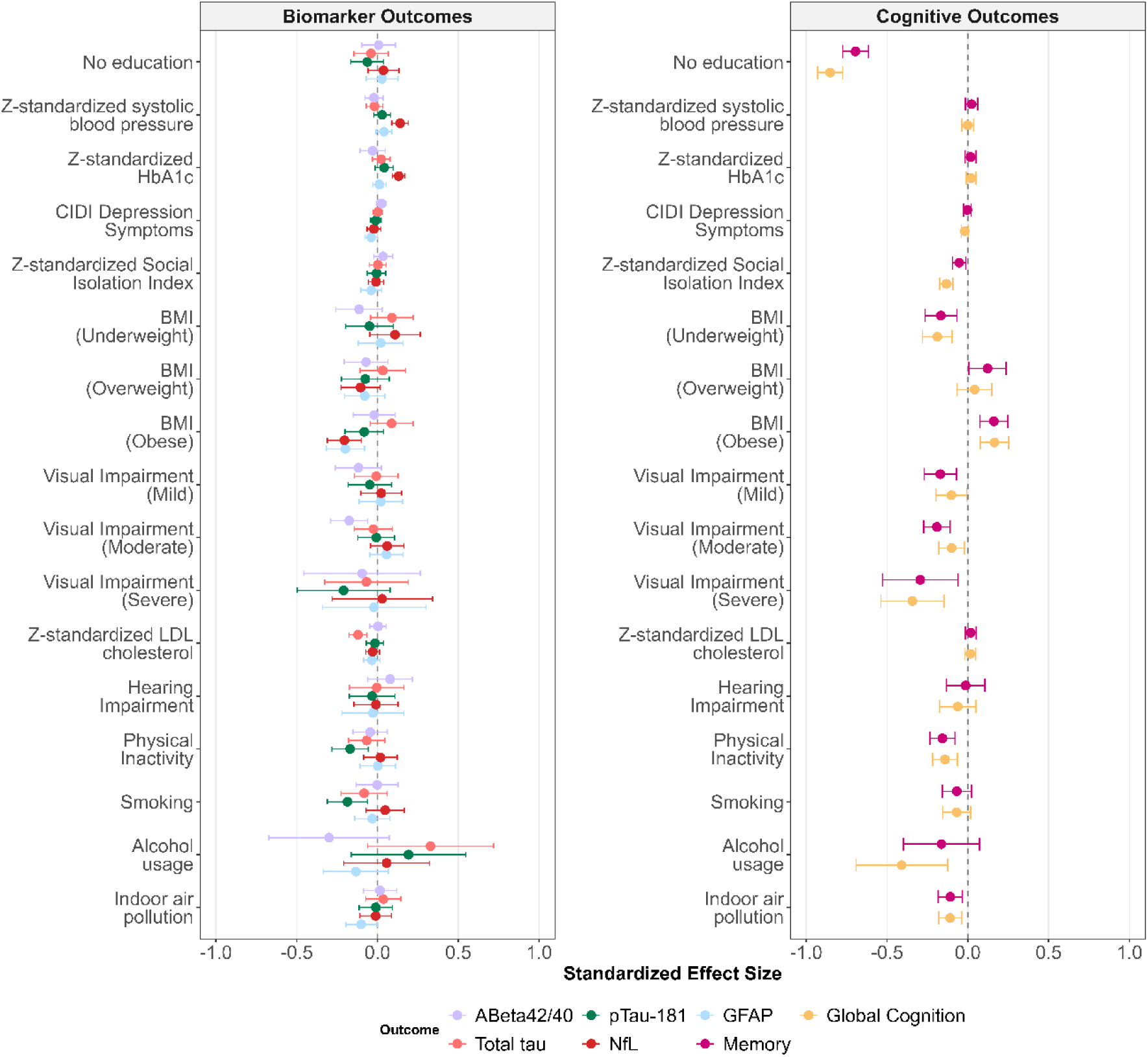
Association of dementia risk factors with cognitive test outcomes and blood-based neurodegenerative biomarkers in LASI-DAD (N=1625). All estimates come from separate linear regression models adjusted for either age, sex/gender, education, household consumption, marital status, and caste (cognitive outcomes) or age, sex/gender, BMI, and eGFR (biomarker outcomes). Error bars show 95% confidence intervals.

Across multiple biomarkers, underweight BMI was associated with biomarker levels more indicative of pathophysiology (Aβ_42/40_ ratio, NfL) and overweight/obese BMI was associated with biomarkers of general neurodegeneration (NfL, GFAP, pTau-181) compared to normal BMI, though only associations between obesity and both NfL and GFAP met the threshold for statistical significance (Figure 1). Additionally, there were several significant associations counter to expectation. Physical inactivity was associated with 0.17SD lower log-transformed p-Tau 181 (p=0.003) and smoking was associated with a 0.19SD lower measure of log-transformed pTau-181 (p=0.003), indicating that individuals with these risk factors had less pTau pathology. Elevated LDL cholesterol was also associated with 0.16SD lower log-transformed total tau (p=0.01), and exposure to indoor air pollution was significantly associated with 0.10SD lower measured GFAP (p=0.04).

Overarching conclusions also held in exploratory models stratified by CDR status, though some heterogeneity was observed. For example, diabetes was significantly associated with higher pTau-181 in the MCI category (β=0.13 SD, p=0.01), but this association was null in the cognitively normal category (Appendix Figure 1).

### Comparisons between findings for cognitive status and biomarker outcomes

Comparisons between findings for biomarker outcomes and findings for cognitive outcomes indicate much smaller standardized effect sizes, a larger proportion of null results, and fewer results in the expected direction for models using biomarker outcomes (Figure 2). For example, having no formal education was significantly associated with lower cognition (β=-0.86SD, p<0.001), but associations with all five biomarkers were null.

**Figure 2.**
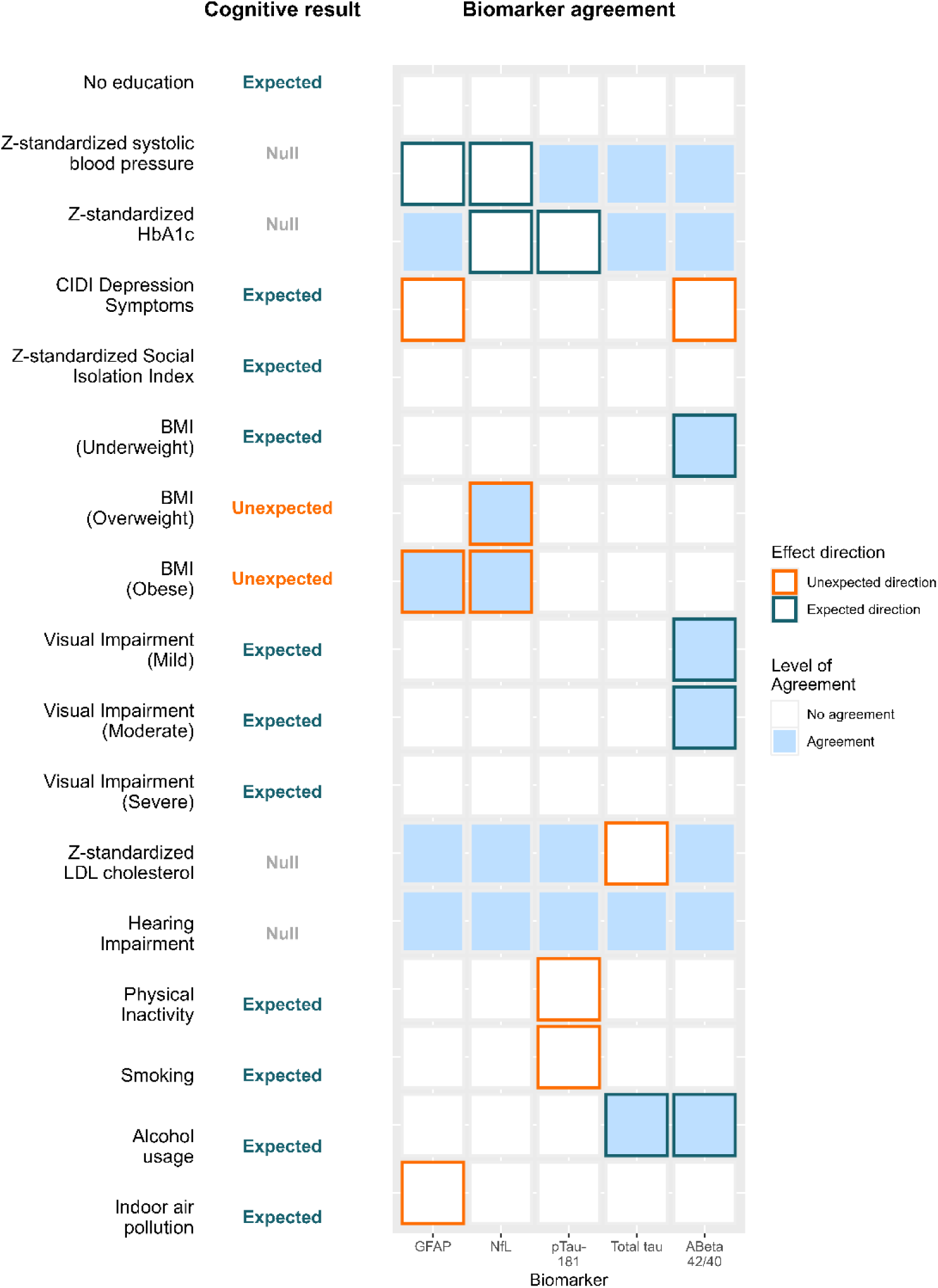
Agreement between cognition test measure and neurodegenerative biomarker results in LASI-DAD (N=1625). α=0.15 is used as a conservative threshold to define significance for agreement and expected/unexpected designations in order to more easily visualize agreement.

Specific areas of disagreement or agreement between the results from models with cognitive outcomes and biomarker outcomes can inform more targeted insights. For example, we replicated cross-sectional analyses that have shown that overweight and obese BMI are associated with higher cognition (inverse to results from many high-income settings),^32^ and results here further illustrate the same pattern was also observed for neurodegenerative biomarkers (NfL and GFAP). Across biomarkers, Aβ42/40 ratio had the largest number of cross-sectional associations in an expected direction and in agreement with results from models for cognitive outcome, with suggestive evidence for associations with alcohol use, underweight BMI, and mild vision impairment, and a significant association with moderate vision impairment (Figure 2). Additionally, while we did not observe statistically significant associations between cardiometabolic risk factors and cognitive outcomes, we did observe cross-sectional associations between NfL and both hypertension and diabetes, and a marginal association between GFAP and hypertension. There were also several more isolated examples of biomarker associations counter to both expectations and results from models with cognitive outcomes (i.e., modifiable risk factors associated with a healthier biomarker profile), including associations between physical inactivity and indoor air pollution and pTau-181.

Our sensitivity analysis suggested that comparisons between results for cognition and biomarker outcomes were not attributable to differences in the adjustment sets (Appendix Figure 2).

## Discussion

In this novel, well-characterized, nationally representative sample of older adults in India, cross-sectional associations between established dementia risk factors and blood-based neurodegenerative markers were largely null, counter to stronger findings with cognitive outcomes for many risk factors. While a few notable exceptions (e.g., NfL and cardiovascular risk factors) may point to specific mechanisms for neurodegeneration, observed inconsistencies largely indicate cross-sectional data from blood-based neurodegenerative biomarkers in isolation may not be associated with dementia risk factors.

Our findings largely correspond with previous work on plasma neurodegenerative biomarkers, which has found some evidence for associations between modifiable dementia risk factors and biomarkers, though results are mixed.^25,26^ Consistent with our finding of associations between NfL and both hypertension and diabetes, the National Health and Nutrition Examination Survey (NHANES), the Framingham Heart Study, and smaller clinical studies have reported that risk factors of cardiovascular disease and diabetes are associated with higher levels of NfL, both in blood and cerebrospinal fluid.^16,33,34^ We also found that vision impairment may be related to lower plasma Aβ_42/40_, and thus increased amyloid pathology in the brain, possibly due to the role of vision impairment as a risk factor for AD^35,36^ or through the effect of AD on vision, via buildup of amyloid proteins in the eye.^37–39^ Prior evidence in other samples has also found that increasing BMI is associated with biomarker levels indicating lower levels of pathology.^40,41^ However, observed associations may be due to potential effects of BMI on the measurement of biomarkers rather than effects on the underlying pathology,^40,41^ for example, if greater blood volume due to higher BMI dilutes biomarker levels in plasma.

Multiple plausible explanations may explain inconsistent and largely null results for associations between dementia risk factors and neurodegenerative biomarkers. The cognitive reserve hypothesis posits that early life exposures, such as education, build resilience through mechanisms that are independent of neuropathology,^42,43^ and may explain why we observed a closer link between some of these risk factors and cognition rather than biomarker outcomes. However, existing research using other forms of biomarker measurement, including imaging, have largely supported the existence of associations between many risk factors — such as cardiovascular disease, vision, and pollution — and brain pathologies.^37,39,44–47^ Alternatively, measurement error may contribute to observed differences, as previous comparisons have shown that blood-based biomarkers may not be as consistent in identifying AD pathologies as CSF or brain-imaging methods, or they may identify pathologies at an earlier disease stage.^12,48^ If the noise present in blood-based neurodegenerative markers outweighs the signal from true effects of risk factors on neurodegenerative pathologies, it may be difficult to detect associations of plasma neurodegenerative biomarkers with risk factors. Additionally, plasma biomarkers may be abnormal earlier in the disease process, and this difference in outcome disease stage may lead to a different association with risk factors measured in later life. We observed some evidence of this heterogeneity in our exploratory CDR-stratified analyses between the cognitively normal and MCI groups, though large uncertainty in estimates makes interpretation of these findings challenging. Given these considerations, for analyses focused on dementia risk factors, plasma neurodegenerative biomarkers may prove more useful when used in conjunction with cognitive outcomes rather than in isolation without consideration of cognitive state. Though plasma biomarkers are a low-cost option in comparison with traditional biomarker modalities, dried blood spots versions of these biomarkers could serve to provide even broader access and lower costs further.

Our study contributes to prior work by analyzing the relationships between dementia risk factors, cognitive test measures, and plasma neurodegenerative biomarkers in an understudied population. The breadth of the survey instrument enabled comparisons across a wide range of dementia risk factors included in the *Lancet* Commission Report. Some limitations should be considered. Though we did include a range of neurodegenerative biomarkers, we could not include markers of vascular neuropathology as plasma-based assays are not currently available. We also included many statistical models and tests, raising concerns about Type I error. Given the large number of models, we also limited our analysis to linear associations, though future research should examine potential non-linearities. However, given the exploratory nature of this present study,^49^ we focused on broader patterns of results to prevent over-interpretation of individual statistical tests that may be due to chance. Our analysis is also cross-sectional, and future work should be done using subsequent waves of LASI-DAD to understand whether patterns hold in longitudinal analyses. The cross-sectional design of this study also means that some risk factors hypothesized to have effects in mid-life were measured in late-life or based on retrospective reports. Furthermore, we were not able to measure ptau217, which should be considered in future studies.

Using cross-sectional data from a nationally representative sample of older adults in India, we found inconsistencies between lifecourse dementia risk factors and associations with cognitive versus blood-based neurodegenerative biomarker outcomes. Associations between lifecourse risk factors and neurodegenerative biomarkers were largely null, though we also identified evidence supporting associations between cardiovascular risk factors and NfL. Overall, findings suggest that blood-based neurodegenerative biomarkers considered in isolation may not be ideal for use in population-based risk factor research, though they remain useful in settings or studies where imaging data are not available and when used in combination with data based on objective cognitive testing. Future research is needed to understand how these biomarkers can be optimally used in conjunction with cognitive testing in large-scale epidemiologic research, for example, by identifying pathological subtypes among those with cognitive impairment or dementia.

## Acknowledgements

We would like to thank the field team, LASI-DAD investigators, and LASI-DAD participants for their time and effort.

## Ethical Considerations

The LASI-DAD study was approved by the University of Southern California IRB on September 28, 2016 (UP-15-00684). All participants gave informed consent for the questionnaire, cognitive testing, and venous blood draw.

## Consent to Participate

All participants provided informed (written or thumbprint) consent for study participation.

## Consent for Publication

Not applicable

## Data availability

Demographic and cognitive assessment data from LASI-DAD are openly available at the Gateway for Global Aging at https://g2aging.org/hrd/get-data. Plasma biomarker data is available upon request from the corresponding author. The data is currently being prepared for public release and will be available at the Gateway for Global Aging shortly.

