## Appendix for "Cross-sectional associations between lifecourse dementia risk factors, plasma neurodegenerative biomarkers, and cognition in older adults in India"

Operationalization of dementia risk factors and other variables

Hypertension was represented in analyses by systolic blood pressure of greater than or equal to 130 mm Hg, or diastolic blood pressure of greater than or equal to 80 mm Hg. Diabetes was represented as greater than or equal to 6.5% hemoglobin A1c (HbA1c) from venous blood samples. High cholesterol was represented using continuous low-density lipoprotein (LDL) of 160 mg/dL or higher from venous blood samples.

Obesity was measured using body mass index (BMI), which was calculated using measured height and weight. Given that associations with BMI may be nonlinear, participants were then categorized into four groups based on their BMI: underweight (<18), normal (18-22.9), overweight (23.0-24.9), and obese (≥25).[50] Vision tests were conducted during point-of-care testing and participants were categorized into four groups based on the distance vision of their better eye: no vision impairment, mild vision impairment, moderate vision impairment, and severe vision impairment/blind. Participants self-reported ever having a hearing or ear-related problem/condition. Depression was measured using the Composite International Diagnostic Interview (CIDI). Social isolation was measured using a twelve-point index which included: playing cards or indoor games, visiting relatives/friends, attending religious functions/events, attending political/community/organization group meetings, not having friends or meeting with friends, marital status, and quality of relationships with household members.

Participants were classified as physically inactive if they responded “hardly ever” or “never” to frequency of moderate or vigorous physical activity. Excessive alcohol consumption was defined as drinking alcohol at least five days per week or binge drinking at least once per week in the past 3 months. Participants were classified as smokers if they self-reported currently smoking or quitting smoking less than one year ago. Indoor air pollution was defined as self-reported use of unclean cooking fuel (kerosene, charcoal, lignite, cola, wood, dung cake).

Self-reported educational attainment was categorized into the following groups: no formal schooling, less than secondary school, and secondary or higher. Self-reported caste was categorized into the following groups: scheduled caste, scheduled tribe, other backward class, and no caste or other caste.

All exposures were measured in late-life or based on retrospective reports from late life (e.g. educational attainment).

Description of LASI-DAD Cognitive Tests and Protocol

As part of the LASI-DAD protocol, respondents were given the following cognitive tests:[22]

Orientation:

- Orientation to time: day of month, month, year, day of week
- Orientation to place: state, city, season, floor of building, area of town/street name, hospital name

Memory:

- Immediate & delayed word recall, word recognition (10 words)
- Immediate & delayed word recall (3 words)
- Immediate & delayed logical memory, logical memory recognition
- Immediate & delayed Brave man story recall
- Constructional praxis delayed recall

Executive functioning:

- Problem solving
- Raven's progressive matrices
- Similarities & differences
- Token test
- Digit span forward and backward
- Go-no-go test
- Symbol cancellation test
- Serial 7s
- Backward day naming

Language/fluency:

- Animal fluency
- Name coconut
- Name scissors
- Name watch
- Name pencil
- Name elbow
- What does one do with a hammer
- Write/say a sentence
- Read and follow a command/follow example

- Phrase repetition
- Where is the local market?
- Follow 3-stage instruction
- Name prime minister

Visuospatial:

- Interlocking pentagons
- Constructional praxis
- Clock drawing

Field workers visited the respondents' homes during the day and used face-to-face Computer-Assisted Personal Interview for the cognitive tests.[22,23] The cognitive tests took 78 minutes on average.[22]

All listed tests were used to develop the factor scores.[24]

Appendix Table 1. Comparison of demographic characteristics between included and excluded participants from the LASI-DAD W1 sample (n=4096)

| **Variable** | | **Mean (SD) or N(%)** |
|---|---|---|
| | **Included (n=1625)** | **Excluded (n=2471)** |
| Age | 68.7 (7.2) | 69.2 (7.8) |
| Gender (% male) | 765 (47.1%) | 1124 (45.5%) |
| Educational attainment | | |
| No schooling | 790 (48.6%) | 1219 (49.3%) |
| Less than secondary | 444 (27.3%) | 632 (25.6%) |
| Secondary or higher | 391 (24.1%) | 620 (25.1%) |
| Married or partnered | 1096 (67.5%) | 1576 (63.8%) |
| Caste | | |
| Scheduled caste | 311 (19.1%) | 438 (17.9%) |
| Scheduled tribe | 62 (3.8%) | 145 (5.9%) |
| Other backward class | 680 (41.9%) | 1059 (43.2%) |
| No caste or other caste | 572 (35.2%) | 808 (33.0%) |
| Per capita consumption quintile | | |
| 1 | 302 (18.5%) | 517 (21.0%) |
| 2 | 354 (21.8%) | 466 (18.9%) |
| 3 | 339 (20.8%) | 480 (19.4%) |
| 4 | 323 (19.9%) | 497 (20.1%) |
| 5 | 308 (19.0%) | 509 (20.6%) |

Appendix Figure 1. Sensitivity analysis of association of dementia risk factors with cognitive test outcomes and blood-based neurodegenerative biomarkers in LASI-DAD (n=1625). All estimates come from separate linear regression models adjusted for either age, sex/gender, education, household consumption, marital status, and caste (cognitive outcomes) or age, sex/gender, education, household consumption, marital status, and caste, BMI, and eGFR (biomarker outcomes). Error bars show 95% confidence intervals.

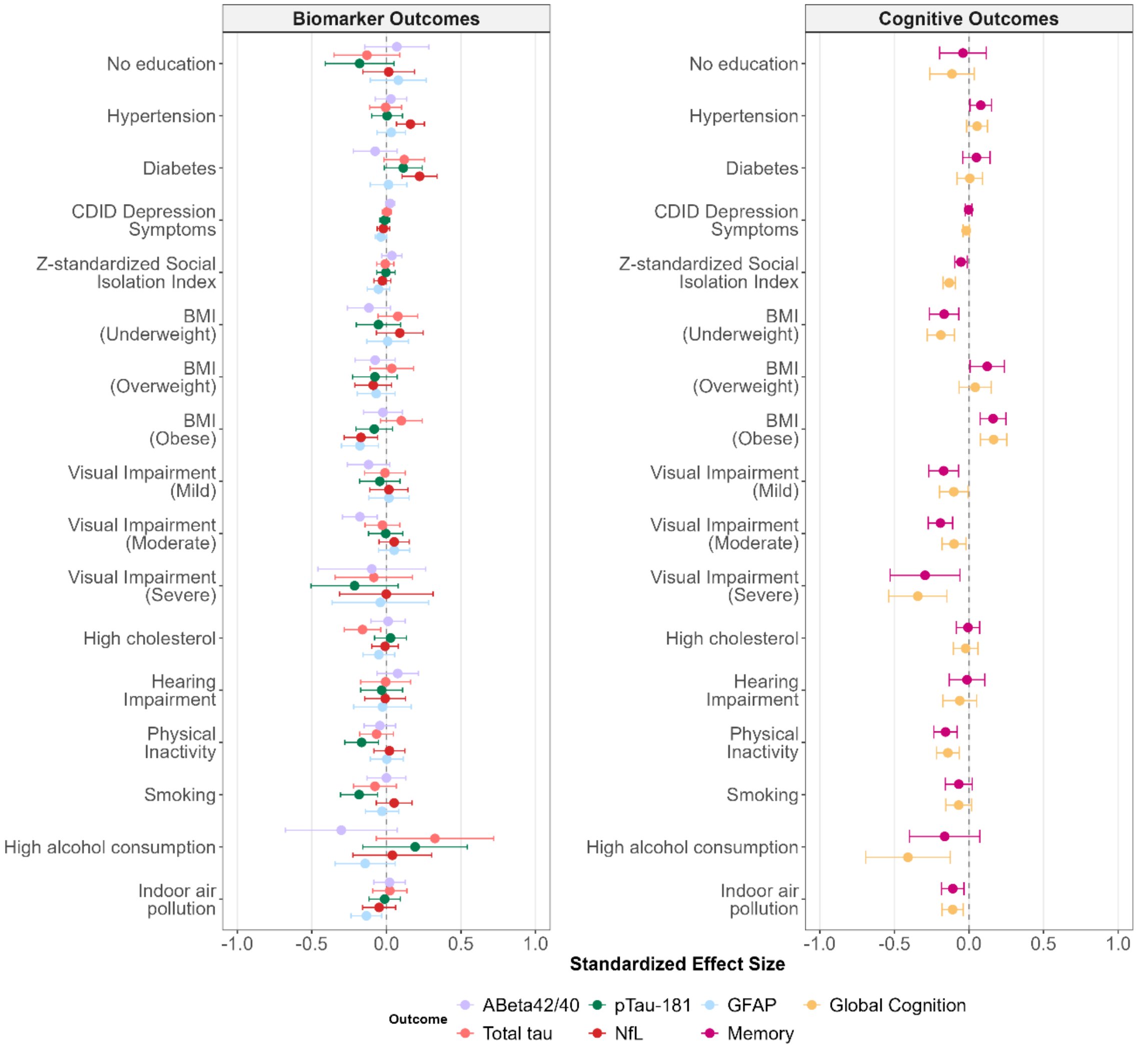

Appendix Figure 2. Sensitivity analysis of association of dementia risk factors with cognitive test outcomes and blood-based neurodegenerative biomarkers stratified by CDR status in LASI-DAD (n=1625). All estimates come from separate linear regression models adjusted for either age, sex/gender, education, household consumption, marital status, and caste (cognitive outcomes) or age, sex/gender, BMI, and eGFR (biomarker outcomes). Error bars show 95% confidence intervals.

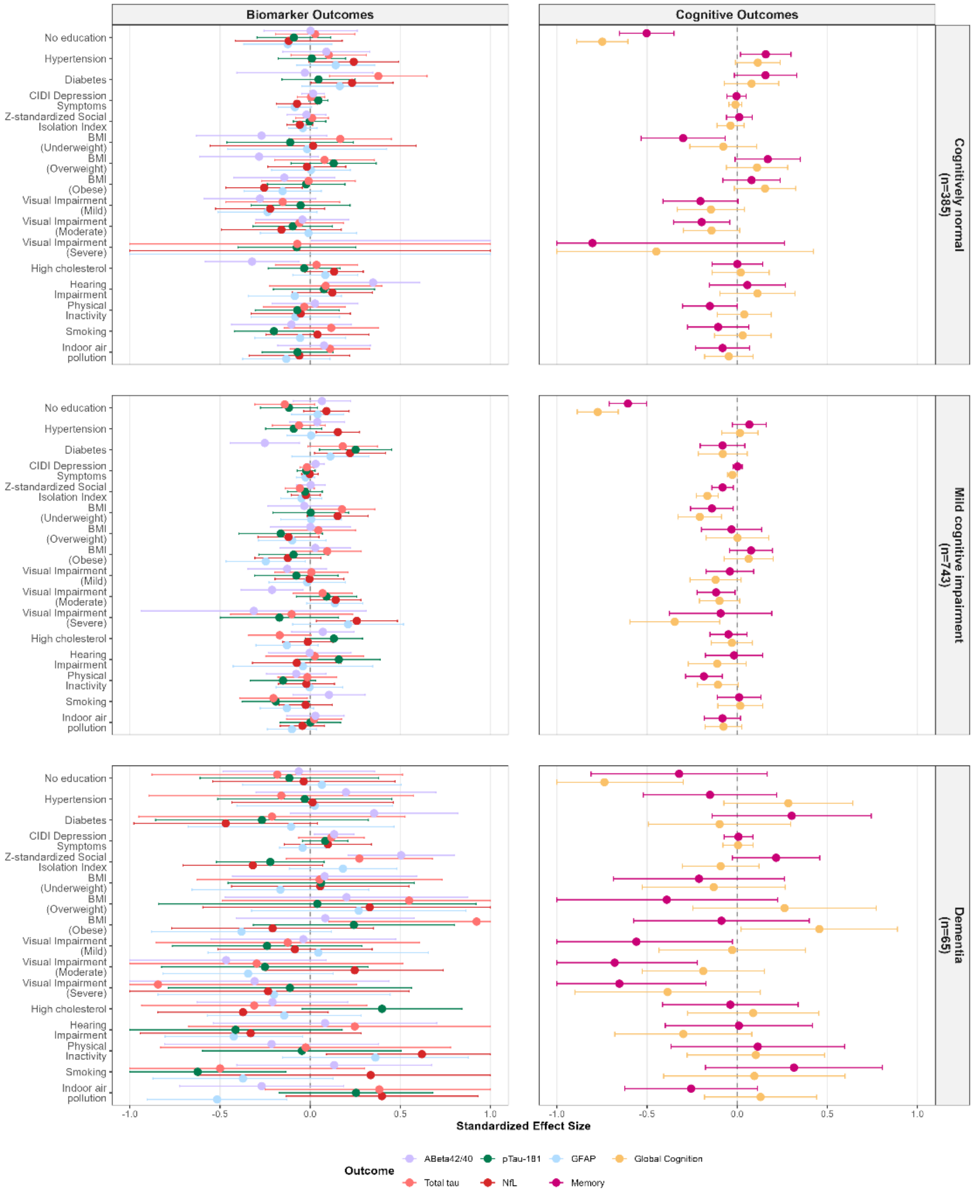
